# Pandemic-Related Post-traumatic Stress Symptomatology in COVID-19 Patients with and without Post-COVID Conditions

**DOI:** 10.1101/2024.01.21.24301574

**Authors:** Emily P. Guinee, Haniya Raza, Elizabeth D. Ballard, Jacob S. Shaw, C. Jason Liang, Michael C. Sneller, Joyce Y. Chung

**Affiliations:** National Institute of Mental Health, Office of the Clinical Director, 10 Center Drive, NIH Building 10, Room 6-5340, Bethesda, MD 20892, USA; National Institute of Mental Health, Experimental Therapeutics & Pathophysiology Branch, NIH Building 10, Room 7-5341, 10 Center Drive, Bethesda, MD 20892; National Institute of Allergy and Infectious Diseases, Biostatistics Research Branch, 5601 Fishers Lane, Rockville MD 20892; National Institute of Allergy and Infectious Diseases, Laboratory of Immunoregulation, 10 Center Drive, NIH Building 10, Room 11C103, Bethesda, MD 20892, USA; Office of Clinical Research Training and Medical Education, National Institutes of Health Clinical Center, 10 Center Drive, Room 1N252E, Bethesda, Maryland 20892

**Keywords:** Post-traumatic stress symptoms, post-traumatic stress disorder, COVID-19, Post-COVID conditions, post-acute sequelae of SARS-CoV-2, resilience

## Abstract

Trauma and stressor-related symptoms have been frequently reported during the COVID-19 pandemic. Few studies compare post-traumatic stress symptoms (PTSS) between patients and non-infected controls. Using data from an ongoing natural history study of COVID-19, this study compared PTSS between patients infected with SARS-CoV-2 during the first year of the pandemic and controls. Within the COVID-19 patient cohort, we also compared PTSS between patients with and without post-COVID conditions, also known as post-acute sequelae of SARS-CoV-2 infection (PASC). This study also examined the association of PTSS with trait resilience and prior trauma exposure. PTSS were assessed using the Impact of Event Scaled-Revised (IES-R), which has a validated probable PTSD cutoff (score ≥33). The results showed that patients (n=131) reported significantly higher IES-R scores than controls (n=82) and had significantly higher odds of having scores indicative of PTSD [AOR: 4.17 *p:* 0.029]. IES-R scores among PASC patients (n=68) were significantly elevated compared to patients without PASC (n=63) and PASC patients did not have higher odds for probable PTSD [AOR: 2.60; *p:* 0.14]. Trait resilience was associated with lower PTSS. These findings help characterize the mental health impact of the COVID-19 illness experience and highlight elevated PTSS in patients with persistent post-COVID conditions.

## Background

In addition to morbidity and mortality associated with severe acute respiratory syndrome coronavirus 2 (SARS-CoV-2) infection, the psychosocial distress related to COVID-19 has contributed to significant mental health burden. In fact, some researchers posit that the COVID-19 pandemic can be understood as a traumatic event (Bridgland et al., 2021). Following exposure to trauma, individuals may develop intrusive thoughts, hyperarousal symptoms, and avoidance behaviors—otherwise known as post-traumatic stress symptoms (PTSS); symptoms that are clinically significant and last for over a month meet criteria for posttraumatic stress disorder (PTSD; (Asssociation, 2013).

Increased incidence of pandemic-related PTSS has been widely reported. Meta-analyses estimates of PTSD pooled prevalence during the pandemic range from 15-22% compared to the World Health Organization’s 2017 global pre-pandemic estimate of 4% (Cénat et al., 2021; Organization, 2017; Yunitri et al., 2022; Zhang et al., 2021). This suggests that pandemic-related stressors have caused significant mental health burden. One such stressor is experiencing COVID-19 infection, which has been found to be associated with higher PTSD prevalence; however, rates remain highly heterogenous and inconsistent (Xie, Xu, & Al-Aly, 2022; Yunitri et al., 2022). While many studies focus on more severely ill hospitalized populations, non-hospitalized COVID-19 patients also experience elevated PTSS following their illness (Einvik et al., 2021; Houben-Wilke et al., 2022; Matsumoto et al., 2022). To date, few studies have compared PTSS between patients previously infected with COVID-19 and non-infected controls (Abdelghani et al., 2021; Matsumoto et al., 2022; Tu et al., 2021; Yuan et al., 2021).

Several factors may impact the risk of PTSS. For example, COVID-19 patients may experience post-COVID conditions, also known as post-acute sequelae of SARS-CoV-2 infection (PASC), which is defined as the persistence of symptoms or complications beyond four weeks after the onset of acute COVID-19 (Nalbandian et al., 2021). Research suggests that patients with PASC may experience more severe PTSS (Houben-Wilke et al., 2022; Matsumoto et al., 2022). In addition to PASC, previous trauma exposure and trait resilience may increase and decrease PTSS, respectively, although their relation to trauma associated with COVID-19 remains underexplored (Bensimon, 2012).

We report findings from a subsample of participants enrolled during the first year of a National Institute of Allergy and Infectious Disease (NIAID) Intramural Research Program protocol (NCT04411147) on the long-term sequelae of COVID-19 (Sneller et al., 2022). Patients with predominantly mild-to-moderate COVID-19 and non-infected controls were enrolled. Published findings from enrollment data identified that female gender, history of anxiety disorder, and current anxiety symptoms were associated with PASC (Sneller et al., 2022). This sub-analysis more closely examines the mental health of the study cohort. Using online surveys and clinical evaluations from participants enrolled during the first year of the study, the aim of this sub-analysis was to compare PTSS severity and probable PTSD prevalence between COVID-19 patients and controls. Within the patient group, we also compared PTSS severity between those with and without PASC. A secondary aim was to examine the association between prior trauma exposure or trait resilience and PTSS.

## Methods

### Participants

Study participants with and without prior SARS-CoV-2 infection from the Mid-Atlantic region were screened and enrolled after clinical evaluation at the National Institutes of Health (NIH) Clinical Center (Sneller et al., 2022). The study was carried out in accordance with the latest version of the Declaration of Helsinki and was approved by the NIH Institutional Review Board. Adults with a confirmed SARS-CoV-2 infection at least six weeks past the onset of symptoms were eligible for the patient group. Controls included adults with no history of SARS-CoV-2 infection and negative testing for antibodies to SARS-CoV-2 nucleocapsid protein. Both study groups were recruited by study posting on ClinicalTrials.gov. Some controls were also recruited if they were close contacts of a patient, though those controls were excluded from this sub-analysis. Controls were not age-nor gender-matched to COVID-19 patients.

### Procedure

Our analysis included participants enrolled between June 30, 2020 and July 1, 2021 (Sneller et al., 2022). All eligible participants provided written informed consent and completed online self-report mental health measures on REDCap (Research Electronic Data Capture) within six weeks of their clinical evaluation, which included a physical exam, laboratory studies, and physiologic measures (Harris et al., 2009). Patients completed surveys a mean of 5.16 months following acute COVID-19 illness.

### Measures

We included online measures of PTSS, prior trauma exposure, trait resilience, mental health treatment history, and current anxiety and depression symptoms. Acute illness severity and PASC status were ascertained at the first study enrollment visit.

### Impact of Event Scale - Revised

COVID-19 pandemic-related PTSS were evaluated using the Impact of Event Scale – Revised (IES-R; (Weiss & Marmar, 1996). This measure screens for PTSS by assessing levels of distress following a traumatic event. We adapted the IES-R prompt to focus on trauma related to the COVID-19 pandemic **(Appendix 1).** Participants reported on specific PTSS experienced in the past seven days. The IES-R has 22 items on a 5-point Likert scale *(0=not at all, 4=extremely),* preceded by an open text field where a participant typed a description of the traumatic event. Item scores are summed for the total score. The IES-R includes three subscales, each assessing a specific symptom cluster of PTSD using DSM-IV criteria: intrusion, avoidance, and hyperarousal (Weiss & Marmar, 1996).

A total score of 33 or greater on the IES-R is a cited cutoff for identifying probable PTSD that would warrant a clinical evaluation for possible PTSD, and thus was used as the cutoff in this analysis (Creamer, Bell, & Failla, 2003). Importantly, IES-R total score is not diagnostic. We also report Impact of Event Scale-6 (IES-6) scores, a briefer set of six items embedded in the IES-R (Thoresen et al., 2010). We included the IES-6 scores to compare our results to surveys that used the IES-6 in larger epidemiological studies during a similar time of the pandemic (Czeisler et al., 2020). Unlike the IES-R, item scores are averaged, and the IES-6 is scored out of 4, with a score of *≥* 1.75 denoting likely PTSD (Hosey et al., 2019).

### Brief Trauma Questionnaire

Prior trauma exposure before COVID-19 was assessed using the Brief Trauma Questionnaire (BTQ; (Schnurr et al., 1999). The BTQ is a ten-item self-report measure that determines if an individual has experienced a significant traumatic event based on Criterion A of the DSM-IV PTSD diagnostic criteria. If participants met a priori significance criteria for one or more traumatic events, based on guidelines from Schnurr and colleagues, they were coded as positive.

### Brief Resilience Scale

The Brief Resilience Scale (BRS) was used to measure trait resilience—the ability to recover from stress (Smith et al., 2008). The BRS is a six-item survey with a 5-point Likert scale. Item scores are totaled, then the average is calculated. BRS scores range from 1-5, with higher scores representing greater resilience.

### Other Mental Health Surveys

Participants also completed a survey on mental health treatment history. If participants endorsed medication use for a mental health condition, mental health hospitalization, or treatment for alcohol and/or drug abuse, they were coded as having a history of mental health treatment. This full measure can be viewed in **Appendix B.** Current depression and anxiety symptoms were measured using the ultra-brief Patient Health Questionnaire-2 (PHQ-2) items and the Generalized Anxiety Disorder-2 (GAD-2) items (Kroenke et al., 2007; Löwe et al., 2010).

### Clinical Measures

Participants were asked about acute illness hospitalization and PASC symptoms at the enrollment clinic visit at the NIH Clinical Center. As defined by the NIAID parent study, PASC is any symptom or medical condition that: 1) began or worsened after the onset of COVID-19 illness or first positive RT-PCR for those who were asymptomatic and 2) was present at the study enrollment visit (Sneller et al., 2022).

### Data Analysis

All analyses were performed using R Statistical Software, version 4.2.2 (R Foundation for Statistical Computing). Unadjusted comparisons of participant demographic and characteristic information were made using the *t* test or Wilcoxon rank-sum test for continuous variables, Fisher exact test for binary variables, and Chi-square test for race.

Many related factors were controlled for in adjusted analyses. The NIAID study identified female gender, self-reported history of anxiety disorder, and current anxiety symptoms as potential risk factors for PASC (Sneller et al., 2022). Age is also implicated in PASC risk (Perlis et al., 2022). Female gender, having a history of psychiatric illness, and race have also been shown to predict PTSS outcomes during the pandemic (Akyuz Cim et al., 2022; Armitage, Dawes, & Munro, 2022; Ashby et al., 2022; Taquet et al., 2021). Accordingly, age, gender, race, mental health treatment history, and current anxiety symptoms (GAD-2) were used as covariates in all adjusted analyses.

IES-R scores by group are reported as medians, rather than means, due to right skew in data distribution (**Supplemental Figure 1-2**). Using the covariates, separate multiple linear regression models were used to predict continuous IES-R scores based on 1) participant group (patient vs control) and 2) patient subgroup (PASC vs. no PASC). Multivariable logistic regression with the same covariates quantified associations on the odds ratio scale between probable PTSD (IES-R *≥ 33, IES-6 ≥ 1.75)* and participant group and patient subgroup, respectively. Using the same covariates, adjusted multiple linear regression models were also used to examine the association of trait resilience (BRS scores) and history of prior trauma (BTQ results) with IES-R scores, in addition to the interaction of these variables with participant group.

## Results

The NIAID study enrolled 189 patients and 120 non-infected control participants between June 30, 2020 and July 1, 2021 (Sneller et al., 2022). Due to a delay in launching online surveys, 131 patients and 82 non-close contact controls who completed the online measures were included in our sub-analysis. The sample was 57% female, 75% White, and ranged from 18 to 78 years old. **Table 1** shows demographic, psychological, and clinical characteristics for patients, controls, and patient subgroups.

**Table 1:** Unadjusted distribution of participant demographic and clinical characteristics across study group status.

|  | Controls (N=82) | COVID-19 Patients (N=131) | COVID-19 PASC (N=68) | COVID-19 No PASC (N=63) |
| --- | --- | --- | --- | --- |
| Mean age (SD, years) | 48.1 (12.5) | 48.7 (13.1) | 48.8 (11.2) | 48.8 (15.3) |
| Female, n (%) | 50 (61.0%) | 72 (55.0%) | 45 (66.2%) | 27 (42.9%) |
| Race, n (%) |  |  |  |  |
| White | 55 (67.1%) | 105 (80.1%) | 54 (79.4%) | 51 (81.0%) |
| Black | 13 (15.9%) | 12 (9.2%) | 5 (7.4%) | 7 (11.1%) |
| Asian | 9 (11.0%) | 8 (6.1%) | 4 (5.9%) | 4 (6.3%) |
| Other* | 5 (6.1%) | 6 (4.6%) | 5 (7.4%) | 1 (1.6%) |
| Hispanic/Latino, n (%) | 19 (17.4%) | 16 (12.2%) | 12 (17.6%) | 4 (6.3%) |
| History of mental health treatment, n (%) | 22 (28.8%) | 36 (27.5%) | 22 (32.3%) | 14 (22.2%) |
| Mean days since COVID diagnosis (SD) | - | 168 (77) | 183 (76) | 153 (75) |
| BTQ positive, n (%) | 43 (52.4%) | 69 (53.4%) | 40 (58.8%) | 29 (46.0%) |
| Mean BRS (SD) | 3.83 (0.70) | 3.86 (0.71) | 3.86 (0.69) | 3.87 (0.74) |
| GAD-2 score $\geq 3$, n (%) | 2 (2.4 %) | 18 (13.7%) | 15 (22.1%) | 3 (4.8%) |
| PHQ-2 score $\geq 3$, n (%) | 2 (2.4%) | 13 (9.9%) | 9 (13.2%) | 4 (6.3%) |
| Hospitalized for COVID-19, n (%) | - | 12 (9.2%) | 5 (7.4%) | 7 (11.1%) |

\* Includes Hawaiian/Pacific Islander, Native Indian/Alaska Native, Mixed Raced, and Unknown
Note: To assess significant group differences, t-tests were performed on means, chi-square test was used for race, and Fisher exact tests assessed all binary variables. Controls and patients significantly differed based on GAD-2 score ( $p=.007$ ). PASC and no PASC patients also significantly varied by GAD-2 score ( $p=.004$ ), in addition to gender ( $p=.009$ ) and mean days since COVID diagnosis ( $p=.024$ ).

### Post-traumatic stress symptomatology (PTSS)

The median IES-R score for the patient group was 11, which was elevated compared to controls (*p =* 0.004). **Table 2** lists median IES-R and IES-6 scores by participant group. Median IES-R and IES-6 scores between patients and controls were significantly different, and this finding was consistent across all three IES-R subscales (**Supplementary Table S1**). Covariate coefficients are reported in **Supplementary Table S2.** Additionally, 4.9% of controls and 18% of patients had IES-R score ≥ 33, denoting probable PTSD. Using a multivariable logistic regression, patients had significantly greater odds of surpassing the IES-R cutoff score compared to controls [adjusted odds ratio (AOR): 4.17; 95% confidence interval (CI): 1.27 – 17.26; *p=*.029], with the IES-6 cutoff (≥ 1.75) yielding similar results (**Table 2**). Additionally, examples of IES-R free responses can be viewed in **Supplemental Table S3**.

**Table 2:** Post-traumatic stress symptom scores and probable PTSD for all study participants.

|  | Controls<br>(N=82) | COVID-19<br>Patients<br>(N=131) | Mean<br>Difference<br>or Adjusted<br>Odds Ratio<br>[95% CI] | *P* | *d*<br>[95% CI] | COVID-19<br>PASC<br>(N=68) | COVID-19<br>No PASC<br>(N=63) | Mean<br>Difference<br>or Adjusted<br>Odds Ratio<br>[95% CI] | *P* | *d*<br>[95% CI] |
| --- | --- | --- | --- | --- | --- | --- | --- | --- | --- | --- |
| Median<br>Total IES-R<br>Score (*IQR*) | 6<br>(1-14.8) | 11<br>(4-25.5) | 5.22<br>[1.66, 8.77] | .004 | 0.48<br>[0.20, 0.75] | 16.5<br>(6.75-33) | 6<br>(2-15) | 6.49<br>[1.7, 11.29] | .009 | 0.67<br>[0.31, 1.02] |
| Median<br>IES-6 Score<br>(*IQR*) | 0.33<br>(0.0-0.8) | 0.67<br>(0.17-1.13) | 0.35<br>[0.14, 0.56] | .001 | 0.54<br>[0.26, 0.82] | 1.17<br>(0.5-2.0) | 0.33<br>(0.0-0.83) | 0.49<br>[0.21-0.77] | <.001 | 0.78<br>[0.42, 1.14] |
| IES-R ≥ 33,<br>*n* (%) | 4<br>(4.9%) | 24<br>(18.3%) | 4.17<br>[1.27, 17.26] | .029 | - | 18<br>(26.5%) | 6<br>(9.5%) | 2.60<br>[0.75, 9.86] | .14 | - |
| IES-6 ≥<br>1.75, *n* (%) | 4<br>(4.9%) | 26<br>(19.8%) | 5.11<br>[1.59, 20.94] | .011 | - | 20<br>(29.4%) | 6<br>(9.5%) | 3.54<br>[1.07, 13.38] | .046 | - |

Note: all analyses were adjusted for gender, race, age, mental health treatment history, and GAD-2.

### Post-acute Sequelae of SARS-CoV-2 (PASC)

Over half of patients (51.9%) reported at least one symptom consistent with PASC. **Table 1** shows the demographic and clinical characteristics of the PASC and no PASC subgroups. **Figure 1** shows the distribution of IES-R scores between controls and patients, with the two patient subgroups specified. Patients with PASC had significantly higher IES-R scores compared to those without PASC (*p=*.01); however when using the IES-R cutoff, the difference between groups did not reach significance [AOR: 2.53; 95% CI: 0.71- 9.77; *p:* 0.157] (**Table 2**). Using the IES-6, PASC patients had both higher IES6 scores and significantly higher odds of meeting the cutoff on the IES-6 [AOR: 3.45; 95% CI: 1.02 – 13.2; *p:* 0.054].

**Figure 1:**
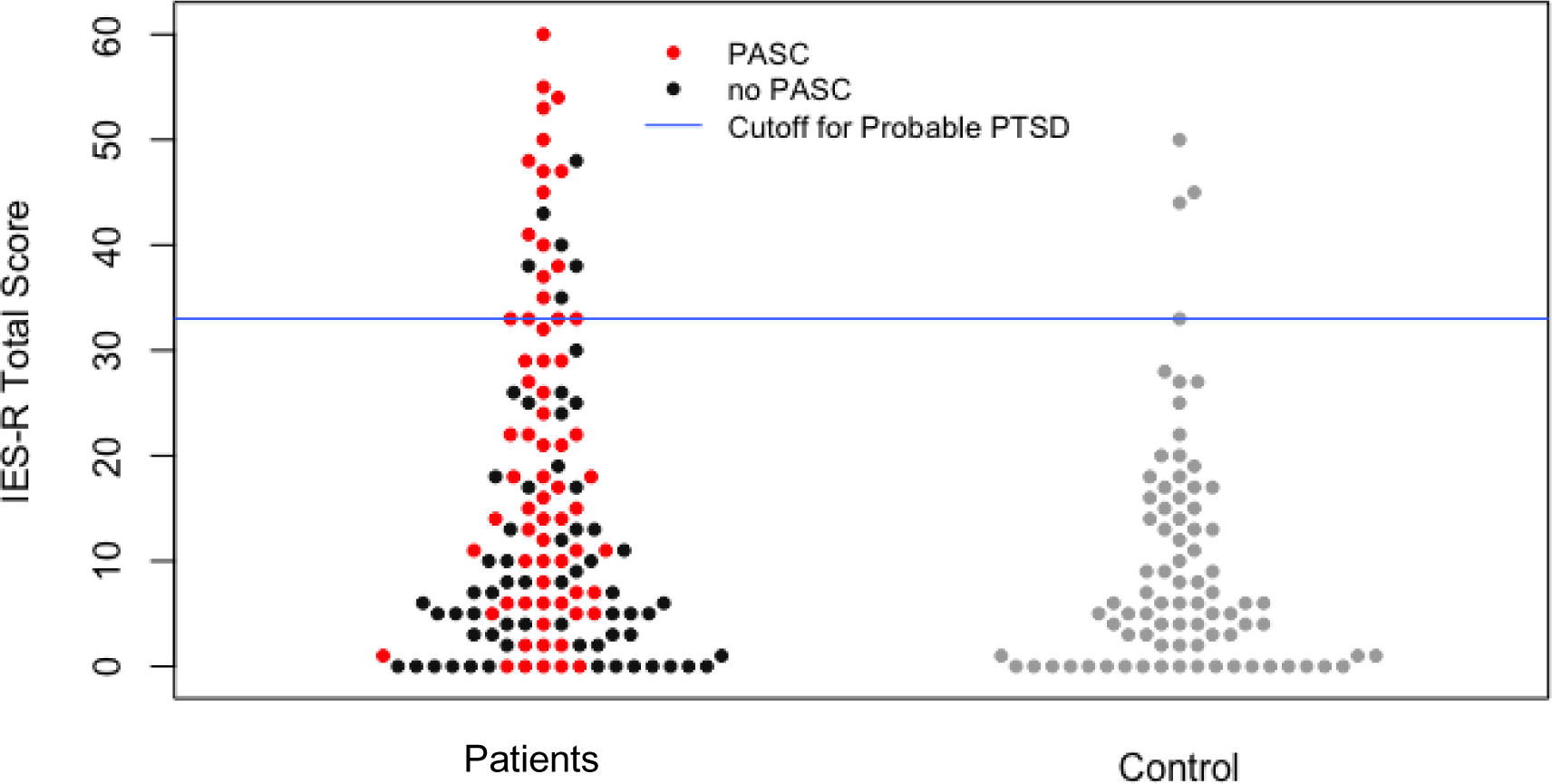
Distribution of IES-R scores among patients (n = 131) with and without PASC and controls (n = 82).

### Trait Resilience & Trauma History

Among the entire participant sample, irrespective of group status, there was a significant main effect of trait resilience (BRS scores) on IES-R scores (β=-3.35 [-6.08, -0.63]; *p=*.004), such that a one-point increase on the BRS (scored from 1 -5) was associated with a mean decrease of 3.35 on the IES-R. The association between traumatic exposure (BTQ positive) and IES-R scores did not reach significance (β=2.68 [-1.21, 6.56]; *p=*.178). There was no significant interaction found between being a patient and either psychological factor.

## Discussion

The objective of the current study was to compare posttraumatic stress symptomatology between COVID-19 patients and controls, and patients with and without PASC. The findings suggest that 1) COVID-19 patients reported elevated PTSS and had significantly greater odds of probable PTSD compared to controls, and more specifically, 2) patients with PASC reported higher PTSS and had greater odds of probable PTSD compared to patients without PASC, though the latter did not reach significance with all measures. Our findings align with previous reports of elevated PTSS and PTSD rates among COVID-19 patients compared to controls in international cohorts (Abdelghani et al., 2021; Matsumoto et al., 2022; Tu et al., 2021; Yuan et al., 2021).

In this study sample, probable PTSD rates in controls were lower than those reported in the general public during the early pandemic (Cénat et al., 2021). For example, a CDC report from late June 2020 using the IES-6 among a U.S. representative sample found 26.3% of respondents with scores *≥ 1.75* (Czeisler et al., 2020). While PTSD prevalence reports among COVID-19 clinical populations range widely from 6.5-50%, we found an 18% rate among our study patients using a survey cutoff score of ≥ 33 (Cénat et al., 2021; Chen et al., 2021; Horn et al., 2020; L. Huang et al., 2022; X. Huang et al., 2022; Janiri et al., 2021; Matsumoto et al., 2022; Tarsitani et al., 2021). The differences among reported estimates could be explained by factors such as study setting and recruitment area, sampling strategy, severity of COVID-19 among patients, methods used for PTSS assessment, and when assessments were done relative to COVID-19 diagnosis.

Over half of our patient sample (51.9%) reported at least one PASC symptom at their baseline visit, which is similar to contemporaneous pooled estimates (Groff et al., 2021; Perlis et al., 2022). PASC patients also reported elevated PTSS when compared to patients without PASC, which is consistent with other reports associating PTSS with post-COVID conditions (L. Huang et al., 2022; Matsumoto et al., 2022). However, in this study these differences were not detected at clinically significant IES-R scores (≥ 33). Our results indicated that self-reported anxiety (GAD-2) was significantly associated with PASC, as was established in the NIAID study (Sneller et al., 2022). It is likely that adjusting for anxiety symptoms (GAD-2), along with small sample size, contributed to why PASC group differences were not detected at more clinically significant PTSS levels (IES-R ≥ 33). Nevertheless, the IES-R findings reported in this sub-study provide more detail on the mental health of patients with and without PASC and controls, since anxiety symptoms (GAD-2) were adjusted for, and IES-R scores still differed substantially among groups. It is possible that trauma or stressor related reactions may better capture the mental health experience of patients with persistent symptoms. However, given the broad definition of PASC, there is much overlap between PASC symptoms and post-traumatic stress symptoms, which confounds the interrelationship of these two variables.

Our study also sought to describe the role of trait resilience and prior trauma exposure in predicting PTSS severity and its association to COVID-19 status. Previous findings indicate that lower resilience is an independent risk factor for PTSD six months after COVID-19 diagnosis, which was supported by our results (Antičević, Bubić, & Britvić, 2021; X. Huang et al., 2022). Trait resilience independently predicted PTSS severity; however, it did not significantly interact with group status. There was no significant association between previous trauma exposure and IES-R scores in our sample. This may have resulted from factors such as sample size, or that a history of prior trauma alone is less predictive of PTSS than peritraumatic processes (Ozer et al., 2003).

Strengths of this study include the use of non-infected controls and validated measures of PTSS, trauma exposure, resilience, and anxiety symptoms. Trauma responses were examined dimensionally (PTSS) and categorically (probable PTSD). To improve construct validity and guide respondents to report on pandemic-related trauma, a pandemic-specific IES-R prompt was utilized. Additionally, many variables associated with PASC and PTSS were controlled for in this analysis, such as anxiety symptoms and gender. Because our study patients had predominantly mild-to-moderate COVID illness, the findings might be more generalizable than other investigations that focus on predominantly hospitalized patient populations.

Several limitations should be acknowledged. The NIAID study enrolled a convenience sample of majority White, non-Hispanic participants, which is not representative of the population most affected by COVID-19 in the U.S. This sample size is rather small, which may have impacted the power to detect some differences. The rate of PASC in the COVID-19 groups may be higher because of self-selection bias (Sneller et al., 2022). Additionally, the IES-R COVID-19-specific prompt could have been interpreted variably by participants (Supplemental Table S2). While the analyses accounted for many potential confounders, days since COVID infection was significantly different between PASC and non-PASC groups and was not adjusted for in analyses.

Moreover, there is debate regarding if the COVID-19 pandemic constitutes a life-threatening Criterion A stressor according to the DSM-5 PTSD diagnostic criteria (Bridgland et al., 2021; Brunet et al., 2022; Van Overmeire, 2020). Perhaps rather than focusing on PTSD alone, other stressor-related disorders, such as adjustment disorder, may provide a better framework for understanding the mental health impact of the pandemic (Brunet et al., 2022). While the IES-R is validated to screen for PTSD, it is not a diagnostic tool and can only suggest likely clinical significance. Future studies should utilize direct clinical evaluations to verify probable PTSD diagnosis rates.

The study was conducted early in the pandemic, when it was unknown how people would experience having COVID-19 and living through a public health crisis. The longitudinal NIAID study will continue to assess trauma-related symptomatology for three years post-enrollment, which will allow ongoing review of clinical findings over time. Overall, our study findings, derived from data collected a mean of 5.16 months following acute COVID-19 illness, suggest that there may be a sustained elevation of PTSS among patients previously infected with SARS-CoV-2, especially those experiencing post-COVID conditions. These findings may guide clinicians to screen their patients for PTSS and stressor-related disorders, particularly among individuals who contracted COVID-19 during the early pandemic and experience PASC.

## Data Availability

Research data are not shared.

## Reactions to COVID related Traumatic Experiences

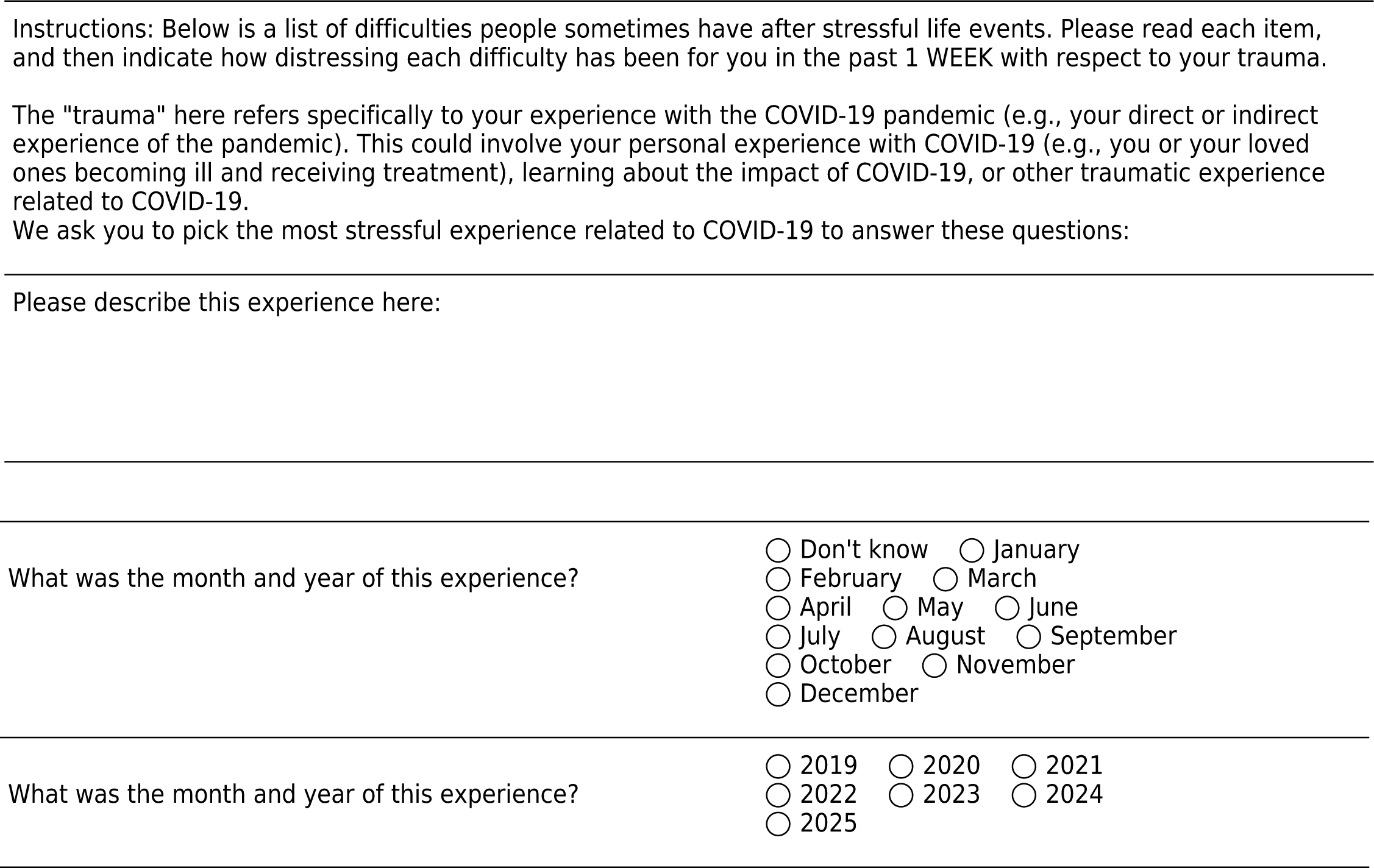
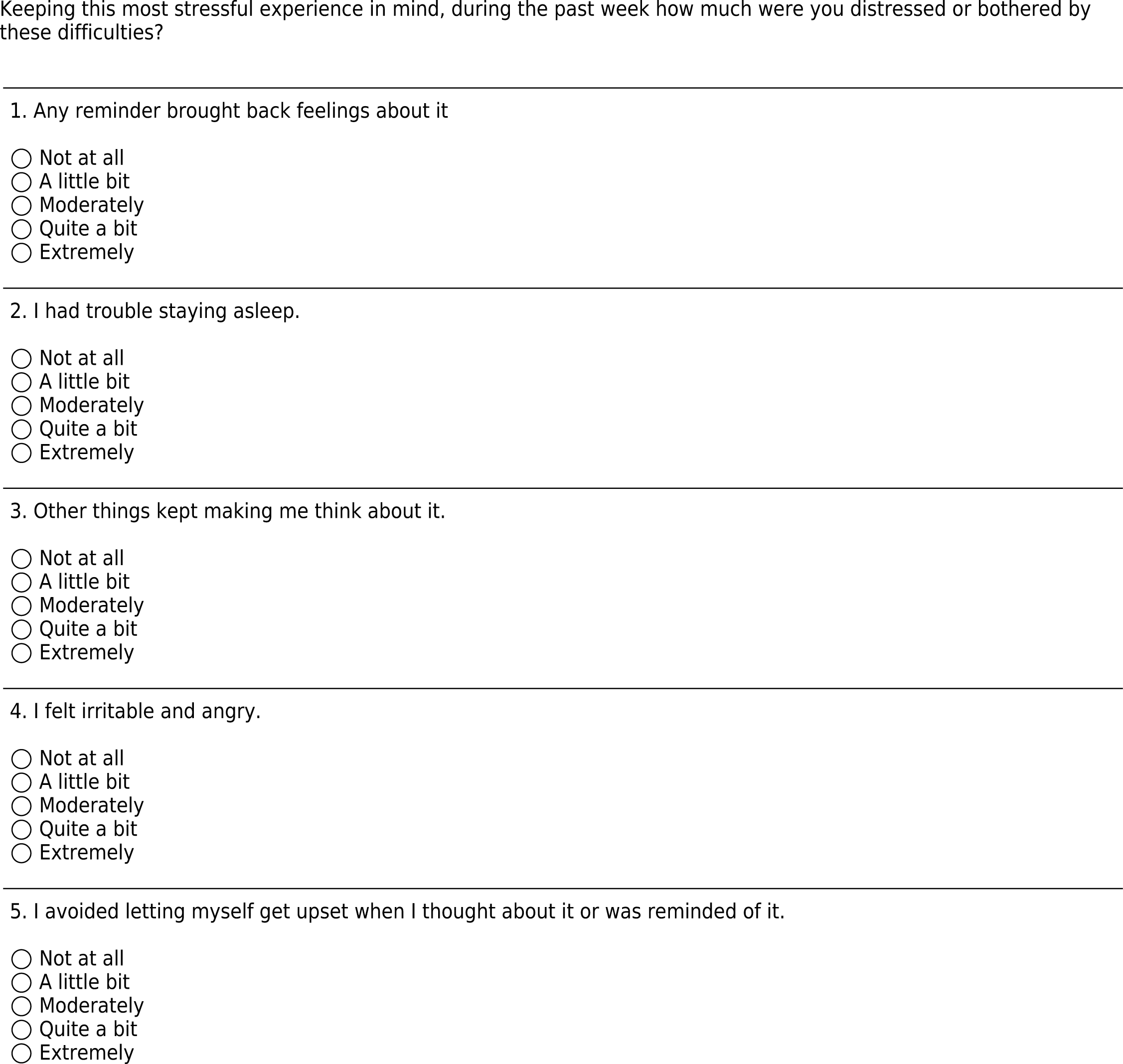
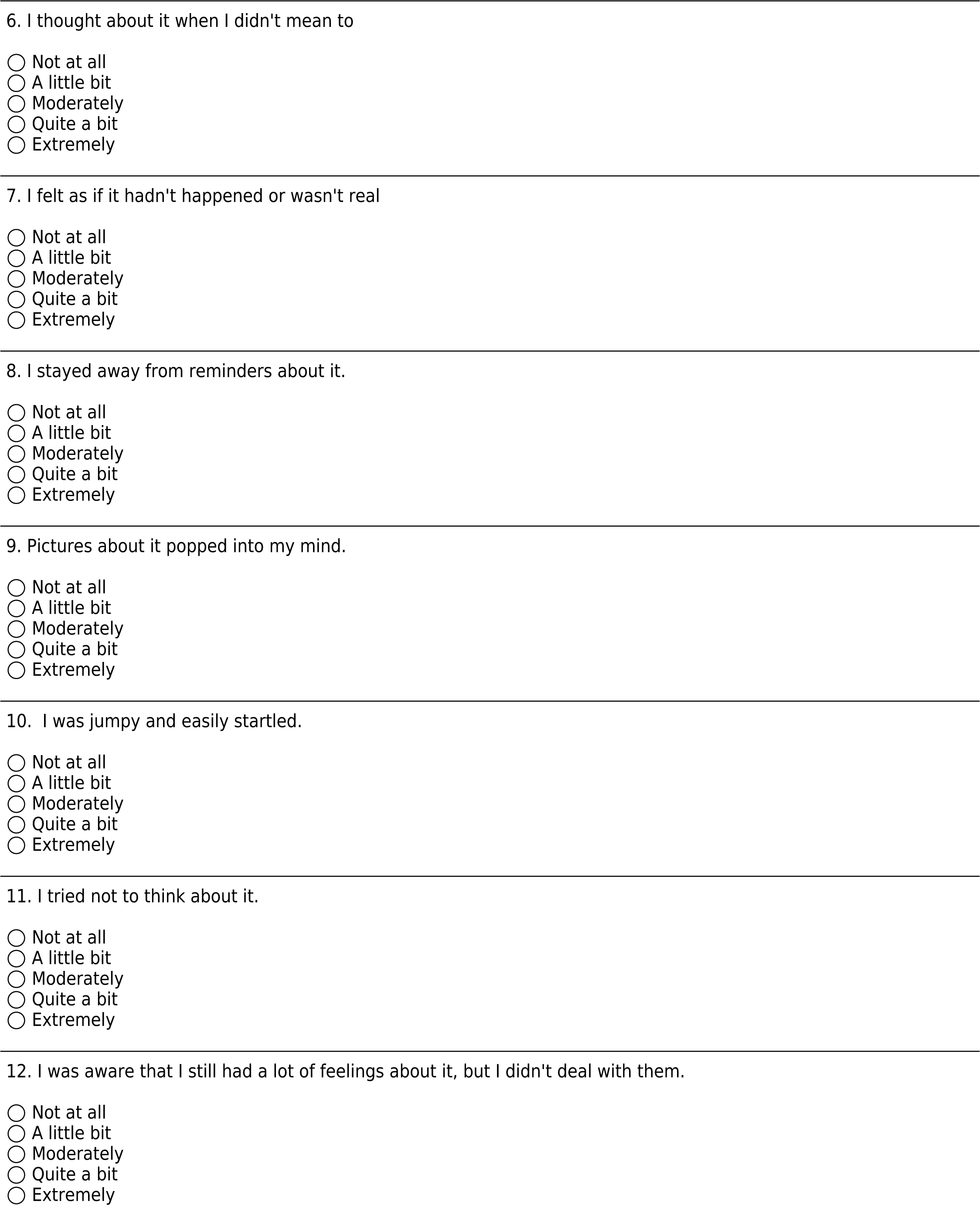
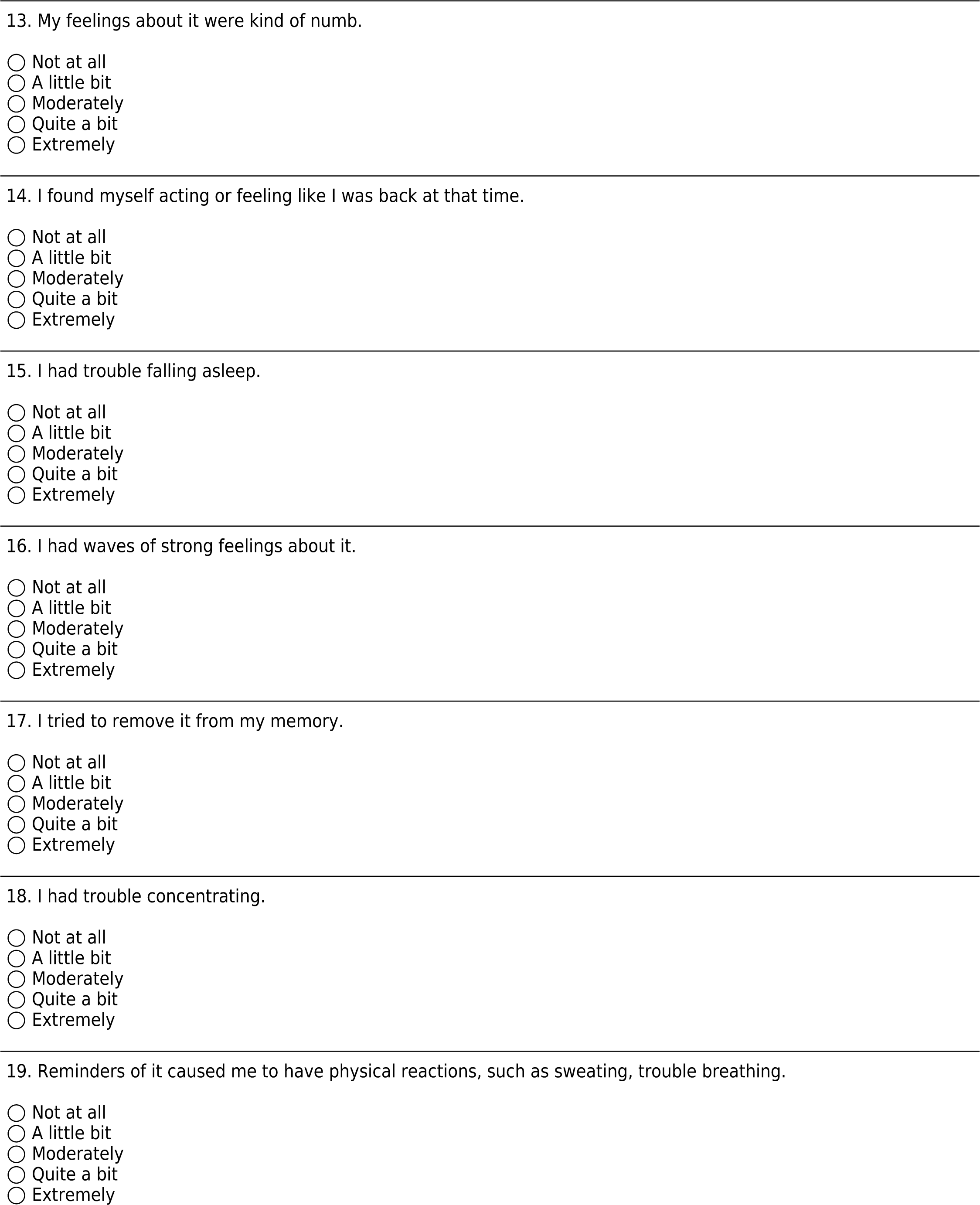
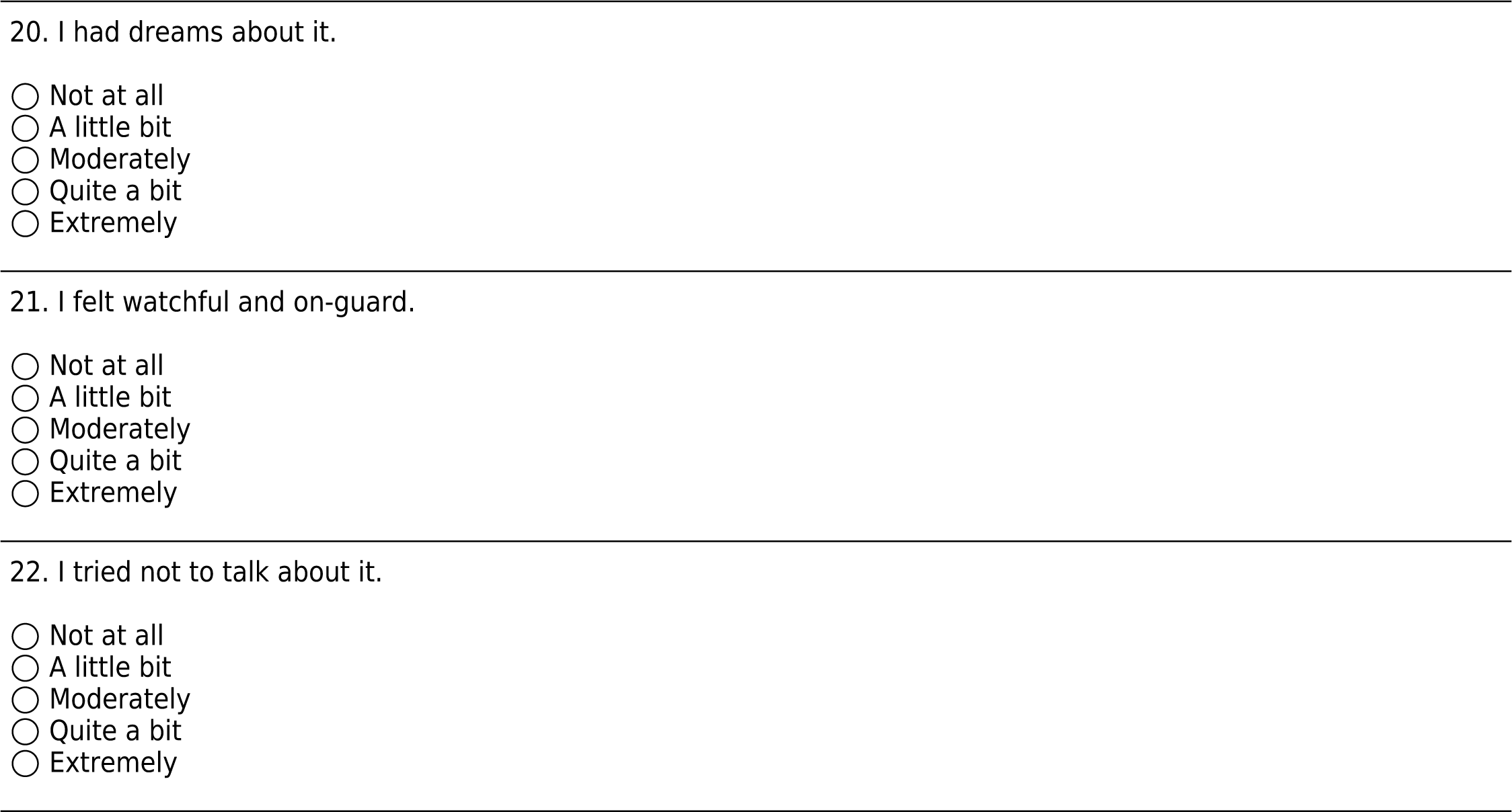

## Mental Health History

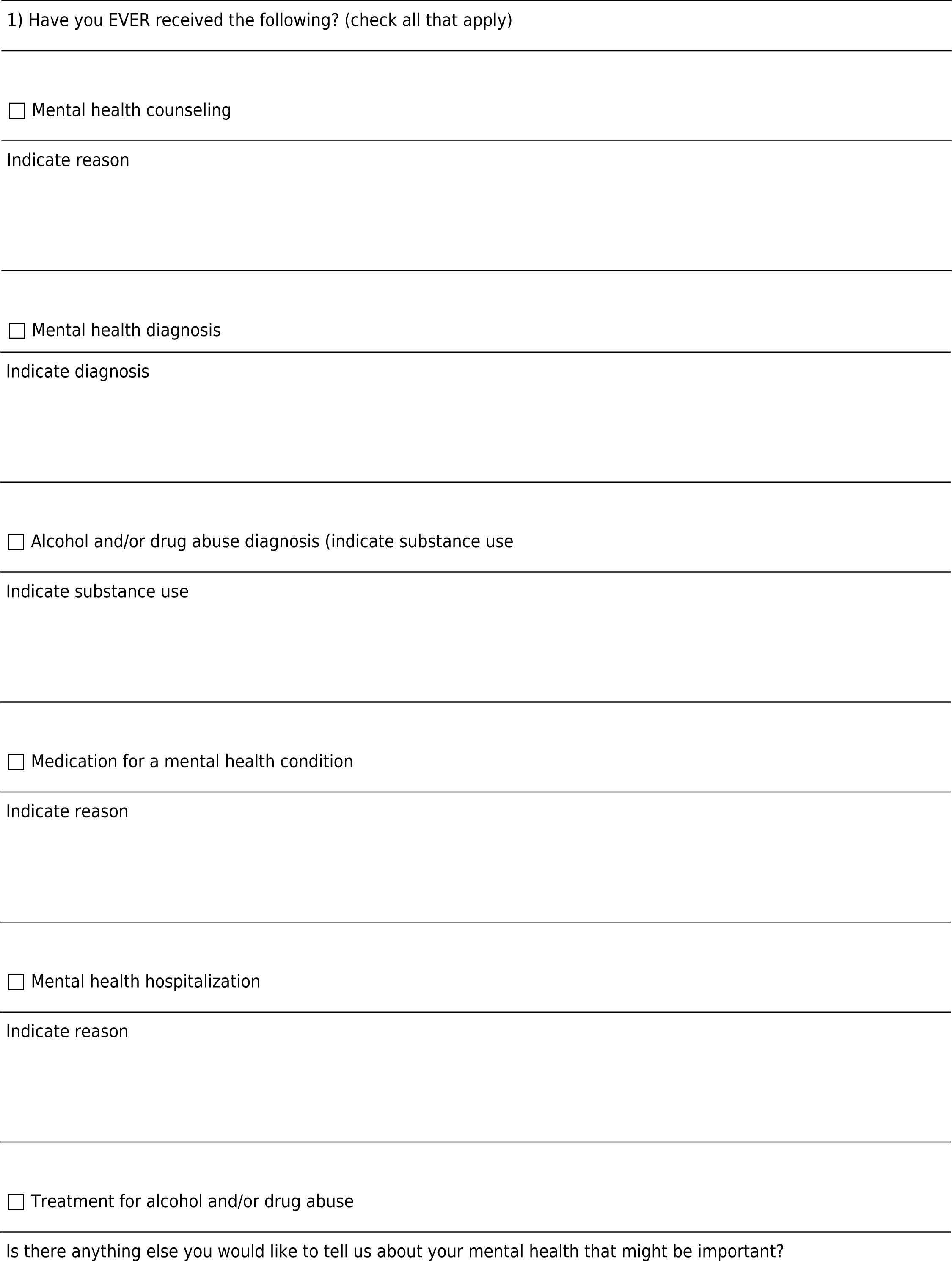

## References

Abdelghani, M., Hassan, M. S., Alsadik, M. E., Abdelmoaty, A. A., Said, A., & Atwa, S. A. (2021). Post-traumatic stress symptoms among an Egyptian sample of post-remission COVID-19 survivors: prevalence and sociodemographic and clinical correlates. Middle East Current Psychiatry, 28(1), 20. 10.1186/s43045-021-00102-y

Akyuz Cim, E. F., Kurhan, F., Dinc, D., & Atli, A. (2022). Assessment of COVID-19 trauma responses. Who has been more traumatized during the pandemic? [Article]. Annales Medico-Psychologiques, 180(6), 503–507. 10.1016/j.amp.2022.01.020

Antičević, V., Bubić, A., & Britvić, D. (2021). Peritraumatic Distress and Posttraumatic Stress Symptoms During the COVID-19 Pandemic: The Contributions of Psychosocial Factors and Pandemic-Related Stressors. Journal of Traumatic Stress, 34(4), 691–700. 10.1002/jts.22701

Armitage, C. J., Dawes, P., & Munro, K. J. (2022). Prevalence and correlates of COVID-19-related traumatic stress symptoms among older adults: A national survey. Journal of Psychiatric Research, 147, 190–193. 10.1016/j.jpsychires.2021.12.054

Ashby, J. S., Rice, K. G., Kira, I. A., & Davari, J. (2022). The relationship of COVID-19 traumatic stress, cumulative trauma, and race to posttraumatic stress disorder symptoms. J Community Psychol, 50(6), 2597–2610. 10.1002/jcop.22762

Asssociation, A. P. (2013). Diagnostic and statistical manual of mental disorders (5th ed.). American Psychiatric Association. 10.1176/appi.books.9780890425596

Bensimon, M. (2012). Elaboration on the association between trauma, PTSD and posttraumatic growth: The role of trait resilience. Personality and Individual Differences, 52(7), 782–787. 10.1016/j.paid.2012.01.011

Bridgland, V. M. E., Moeck, E. K., Green, D. M., Swain, T. L., Nayda, D. M., Matson, L. A., Hutchison, N. P., & Takarangi, M. K. T. (2021). Why the COVID-19 pandemic is a traumatic stressor. PLoS ONE, 16(1), e0240146. 10.1371/journal.pone.0240146

Brunet, A., Rivest-Beauregard, M., Lonergan, M., Cipolletta, S., Rasmussen, A., Meng, X., Jaafari, N., Romero, S., Superka, J., Brown, A. D., & Sapkota, R. P. (2022). PTSD is not the emblematic disorder of the COVID-19 pandemic; adjustment disorder is. BMC Psychiatry, 22(1), 300. 10.1186/s12888-022-03903-5

Cénat, J. M., Blais-Rochette, C., Kokou-Kpolou, C. K., Noorishad, P. G., Mukunzi, J. N., McIntee, S. E., Dalexis, R. D., Goulet, M. A., & Labelle, P. R. (2021). Prevalence of symptoms of depression, anxiety, insomnia, posttraumatic stress disorder, and psychological distress among populations affected by the COVID-19 pandemic: A systematic review and meta-analysis. Psychiatry Res, 295, 113599. 10.1016/j.psychres.2020.113599

Chen, Y., Huang, X., Zhang, C., An, Y., Liang, Y., Yang, Y., & Liu, Z. (2021). Prevalence and predictors of posttraumatic stress disorder, depression and anxiety among hospitalized patients with coronavirus disease 2019 in China. BMC Psychiatry, 21(1), 80. 10.1186/s12888-021-03076-7

Creamer, M., Bell, R., & Failla, S. (2003). Psychometric properties of the Impact of Event Scale - Revised. Behav Res Ther, 41(12), 1489–1496. 10.1016/j.brat.2003.07.010

Czeisler, M., Lane, R. I., Petrosky, E., Wiley, J. F., Christensen, A., Njai, R., Weaver, M. D., Robbins, R., Facer-Childs, E. R., Barger, L. K., Czeisler, C. A., Howard, M. E., & Rajaratnam, S. M. W. (2020). Mental Health, Substance Use, and Suicidal Ideation During the COVID-19 Pandemic - United States, June 24-30, 2020. MMWR Morb Mortal Wkly Rep, 69(32), 1049–1057. 10.15585/mmwr.mm6932a1

Einvik, G., Dammen, T., Ghanima, W., Heir, T., & Stavem, K. (2021). Prevalence and Risk Factors for Post-Traumatic Stress in Hospitalized and Non-Hospitalized COVID-19 Patients. Int J Environ Res Public Health, 18(4). 10.3390/ijerph18042079

Groff, D., Sun, A., Ssentongo, A. E., Ba, D. M., Parsons, N., Poudel, G. R., Lekoubou, A., Oh, J. S., Ericson, J. E., Ssentongo, P., & Chinchilli, V. M. (2021). Short-term and Long-term Rates of Postacute Sequelae of SARS-CoV-2 Infection: A Systematic Review. JAMA Network Open, 4(10), e2128568–e2128568. 10.1001/jamanetworkopen.2021.28568

Harris, P. A., Taylor, R., Thielke, R., Payne, J., Gonzalez, N., & Conde, J. G. (2009). Research electronic data capture (REDCap)--a metadata-driven methodology and workflow process for providing translational research informatics support. J Biomed Inform, 42(2), 377–381. 10.1016/j.jbi.2008.08.010

Horn, M., Wathelet, M., Fovet, T., Amad, A., Vuotto, F., Faure, K., Astier, T., Noël, H., Duhem, S., Vaiva, G., D’Hondt, F., & Henry, M. (2020). Is COVID-19 Associated With Posttraumatic Stress Disorder? J Clin Psychiatry, 82(1). 10.4088/JCP.20m13641

Hosey, M. M., Leoutsakos, J.-M. S., Li, X., Dinglas, V. D., Bienvenu, O. J., Parker, A. M., Hopkins, R. O., Needham, D. M., & Neufeld, K. J. (2019). Screening for posttraumatic stress disorder in ARDS survivors: validation of the Impact of Event Scale-6 (IES-6). Critical Care, 23(1), 276. 10.1186/s13054-019-2553-z

Houben-Wilke, S., Goërtz, Y. M., Delbressine, J. M., Vaes, A. W., Meys, R., Machado, F. V., van Herck, M., Burtin, C., Posthuma, R., Franssen, F. M., Vijlbrief, H., Spies, Y., van ’t Hul, A. J., Spruit, M. A., & Janssen, D. J. (2022). The Impact of Long COVID-19 on Mental Health: Observational 6-Month Follow-Up Study. JMIR Ment Health, 9(2), e33704. 10.2196/33704

Huang, L., Xu, X., Zhang, L., Zheng, D., Liu, Y., Feng, B., Hu, J., Lin, Q., Xi, X., Wang, Q., Lin, M., Zhou, X., He, Z., Weng, H., Deng, Q., Ding, B., Guo, J., & Zhang, Z. (2022). Post-traumatic Stress Disorder Symptoms and Quality of Life of COVID-19 Survivors at 6-Month Follow-Up: A Cross-Sectional Observational Study [Original Research]. Frontiers in Psychiatry, 12. 10.3389/fpsyt.2021.782478

Huang, X., Liu, L., Eli, B., Wang, J., Chen, Y., & Liu, Z. (2022). Mental Health of COVID-19 Survivors at 6 and 12 Months Postdiagnosis: A Cohort Study [Article]. Frontiers in Psychiatry, 13. 10.3389/fpsyt.2022.863698

Janiri, D., Carfì, A., Kotzalidis, G. D., Bernabei, R., Landi, F., Sani, G., & Group, G. A. C.-P.-A. C. S. (2021). Posttraumatic Stress Disorder in Patients After Severe COVID-19 Infection. JAMA Psychiatry, 78(5), 567–569. 10.1001/jamapsychiatry.2021.0109

Kroenke, K., Spitzer, R. L., Williams, J. B., Monahan, P. O., & Löwe, B. (2007). Anxiety disorders in primary care: prevalence, impairment, comorbidity, and detection. Ann Intern Med, 146(5), 317–325. 10.7326/0003-4819-146-5-200703060-00004

Löwe, B., Wahl, I., Rose, M., Spitzer, C., Glaesmer, H., Wingenfeld, K., Schneider, A., & Brähler, E. (2010). A 4-item measure of depression and anxiety: Validation and standardization of the Patient Health Questionnaire-4 (PHQ-4) in the general population. Journal of Affective Disorders, 122(1), 86–95. 10.1016/j.jad.2009.06.019

Matsumoto, K., Hamatani, S., Shimizu, E., Käll, A., & Andersson, G. (2022). Impact of post-COVID conditions on mental health: a cross-sectional study in Japan and Sweden. BMC Psychiatry, 22(1), 237. 10.1186/s12888-022-03874-7

Nalbandian, A., Sehgal, K., Gupta, A., Madhavan, M. V., McGroder, C., Stevens, J. S., Cook, J. R., Nordvig, A. S., Shalev, D., Sehrawat, T. S., Ahluwalia, N., Bikdeli, B., Dietz, D., Der-Nigoghossian, C., Liyanage-Don, N., Rosner, G. F., Bernstein, E. J., Mohan, S., Beckley, A. A., . . . Wan, E. Y. (2021). Post-acute COVID-19 syndrome. Nature Medicine, 27(4), 601–615. 10.1038/s41591-021-01283-z

Organization, W. H. (2017). Depression and other common mental disorders: global health estimates.

Ozer, E. J., Best, S. R., Lipsey, T. L., & Weiss, D. S. (2003). Predictors of posttraumatic stress disorder and symptoms in adults: a meta-analysis. Psychol Bull, 129(1), 52–73. 10.1037/0033-2909.129.1.52

Perlis, R. H., Santillana, M., Ognyanova, K., Safarpour, A., Lunz Trujillo, K., Simonson, M. D., Green, J., Quintana, A., Druckman, J., Baum, M. A., & Lazer, D. (2022). Prevalence and Correlates of Long COVID Symptoms Among US Adults. JAMA Network Open, 5(10), e2238804–e2238804. 10.1001/jamanetworkopen.2022.38804

Schnurr, P., Vielhauer, M., Weathers, F., & Findler, M. (1999). The Brief Trauma Questionnaire (BTQ).

Smith, B. W., Dalen, J., Wiggins, K., Tooley, E., Christopher, P., & Bernard, J. (2008). The brief resilience scale: assessing the ability to bounce back. Int J Behav Med, 15(3), 194–200. 10.1080/10705500802222972

Sneller, M. C., Liang, C. J., Marques, A. R., Chung, J. Y., Shanbhag, S. M., Fontana, J. R., Raza, H., Okeke, O., Dewar, R. L., Higgins, B. P., Tolstenko, K., Kwan, R. W., Gittens, K. R., Seamon, C. A., McCormack, G., Shaw, J. S., Okpali, G. M., Law, M., Trihemasava, K., . . . Lane, H. C. (2022). A Longitudinal Study of COVID-19 Sequelae and Immunity: Baseline Findings. Annals of Internal Medicine, 175(7), 969–979. 10.7326/M21-4905

Taquet, M., Luciano, S., Geddes, J. R., & Harrison, P. J. (2021). Bidirectional associations between COVID-19 and psychiatric disorder: retrospective cohort studies of 62&#x2008;354 COVID-19 cases in the USA. The Lancet Psychiatry, 8(2), 130–140. 10.1016/S2215-0366(20)30462-4

Tarsitani, L., Vassalini, P., Koukopoulos, A., Borrazzo, C., Alessi, F., Di Nicolantonio, C., Serra, R., Alessandri, F., Ceccarelli, G., Mastroianni, C. M., & d’Ettorre, G. (2021). Post-traumatic Stress Disorder Among COVID-19 Survivors at 3-Month Follow-up After Hospital Discharge. Journal of General Internal Medicine, 36(6), 1702–1707. 10.1007/s11606-021-06731-7

Thoresen, S., Tambs, K., Hussain, A., Heir, T., Johansen, V. A., & Bisson, J. I. (2010). Brief measure of posttraumatic stress reactions: impact of Event Scale-6. Soc Psychiatry Psychiatr Epidemiol, 45(3), 405–412. 10.1007/s00127-009-0073-x

Tu, Y., Zhang, Y., Li, Y., Zhao, Q., Bi, Y., Lu, X., Kong, Y., Wang, L., Lu, Z., & Hu, L. (2021). Post-traumatic stress symptoms in COVID-19 survivors: a self-report and brain imaging follow-up study. Mol Psychiatry, 26(12), 7475–7480. 10.1038/s41380-021-01223-w

Van Overmeire, R. (2020). The Methodological Problem of Identifying Criterion A Traumatic Events During the COVID-19 Era: A Commentary on Karatzias et al. (2020). Journal of Traumatic Stress, 33(5), 864-865. 10.1002/jts.22594

Weiss, D. S., & Marmar, C. R. (1996). The Impact of Event Scale - Revised. In J. K. Wilson, T.M. (Ed.), Assessing psychological trauma and PTSD (pp. 399–411). Guilford.

Xie, Y., Xu, E., & Al-Aly, Z. (2022). Risks of mental health outcomes in people with covid-19: cohort study. *BMJ*, e068993. 10.1136/bmj-2021-068993

Yuan, Y., Liu, Z. H., Zhao, Y. J., Zhang, Q., Zhang, L., Cheung, T., Jackson, T., Jiang, G. Q., & Xiang, Y. T. (2021). Prevalence of Post-traumatic Stress Symptoms and Its Associations With Quality of Life, Demographic and Clinical Characteristics in COVID-19 Survivors During the Post-COVID-19 Era. Front Psychiatry, 12, 665507. 10.3389/fpsyt.2021.665507

Yunitri, N., Chu, H., Kang, X. L., Jen, H. J., Pien, L. C., Tsai, H. T., Kamil, A. R., & Chou, K. R. (2022). Global prevalence and associated risk factors of posttraumatic stress disorder during COVID-19 pandemic: A meta-analysis. Int J Nurs Stud, 126, 104136. 10.1016/j.ijnurstu.2021.104136

Zhang, L., Pan, R., Cai, Y., & Pan, J. (2021). The Prevalence of Post-Traumatic Stress Disorder in the General Population during the COVID-19 Pandemic: A Systematic Review and Single-Arm Meta-Analysis. Psychiatry Investig, 18(5), 426–433. 10.30773/pi.2020.0458

