## Supplemental Figures for "Pandemic-Related Post-traumatic Stress Symptomatology in COVID-19 Patients with and without Post-COVID Conditions"

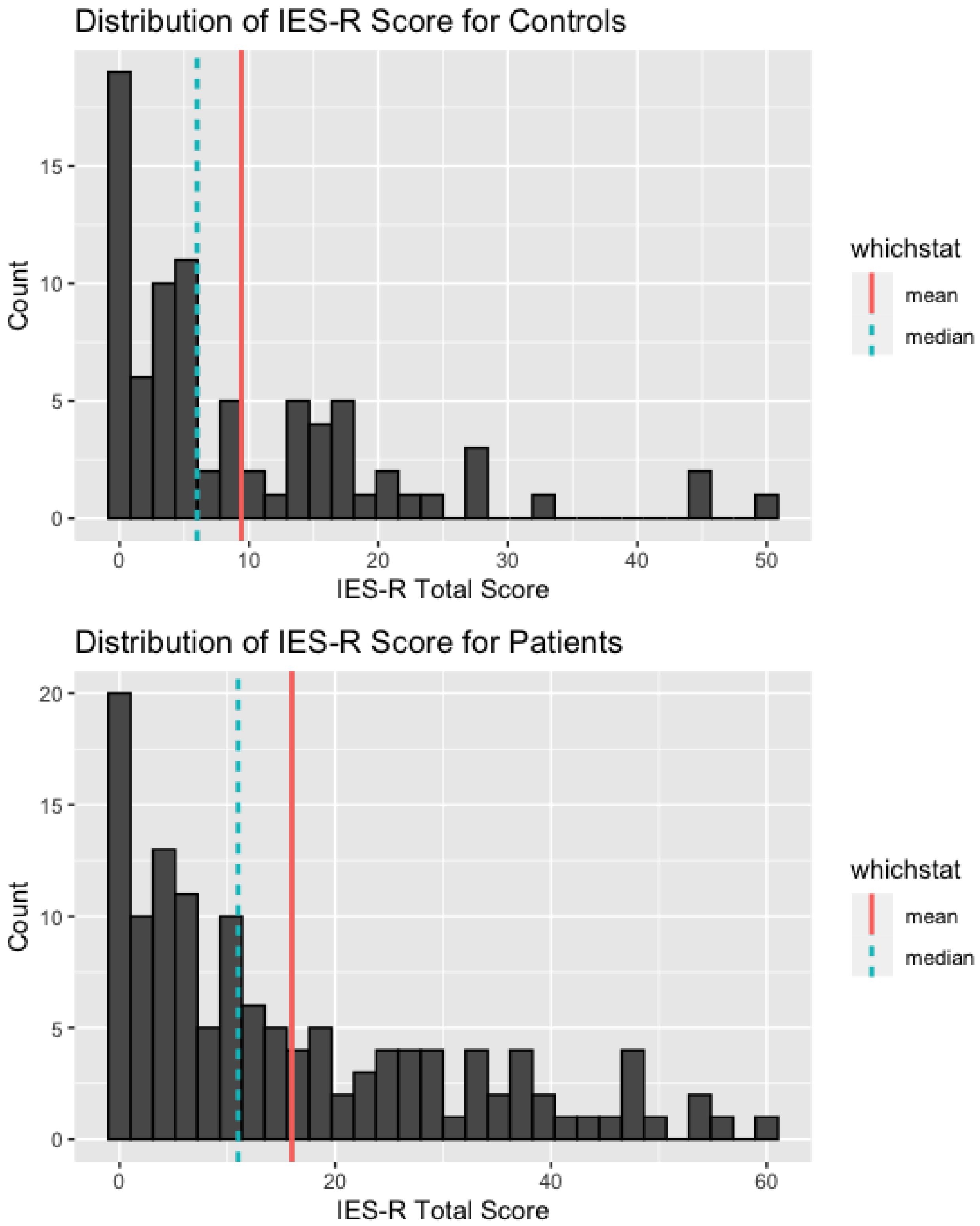


Supplementary Figure 1: Histogram of IES-R score distribution displaying skew in data. Among controls, mean IES-R is 9.41 (SD=10.83) and median is 6 (*IQR*=1.0-14.75). Among patients, mean score is 15.98 (SD=15.35) and median is 11 (*IQR*=4.0-25.0).

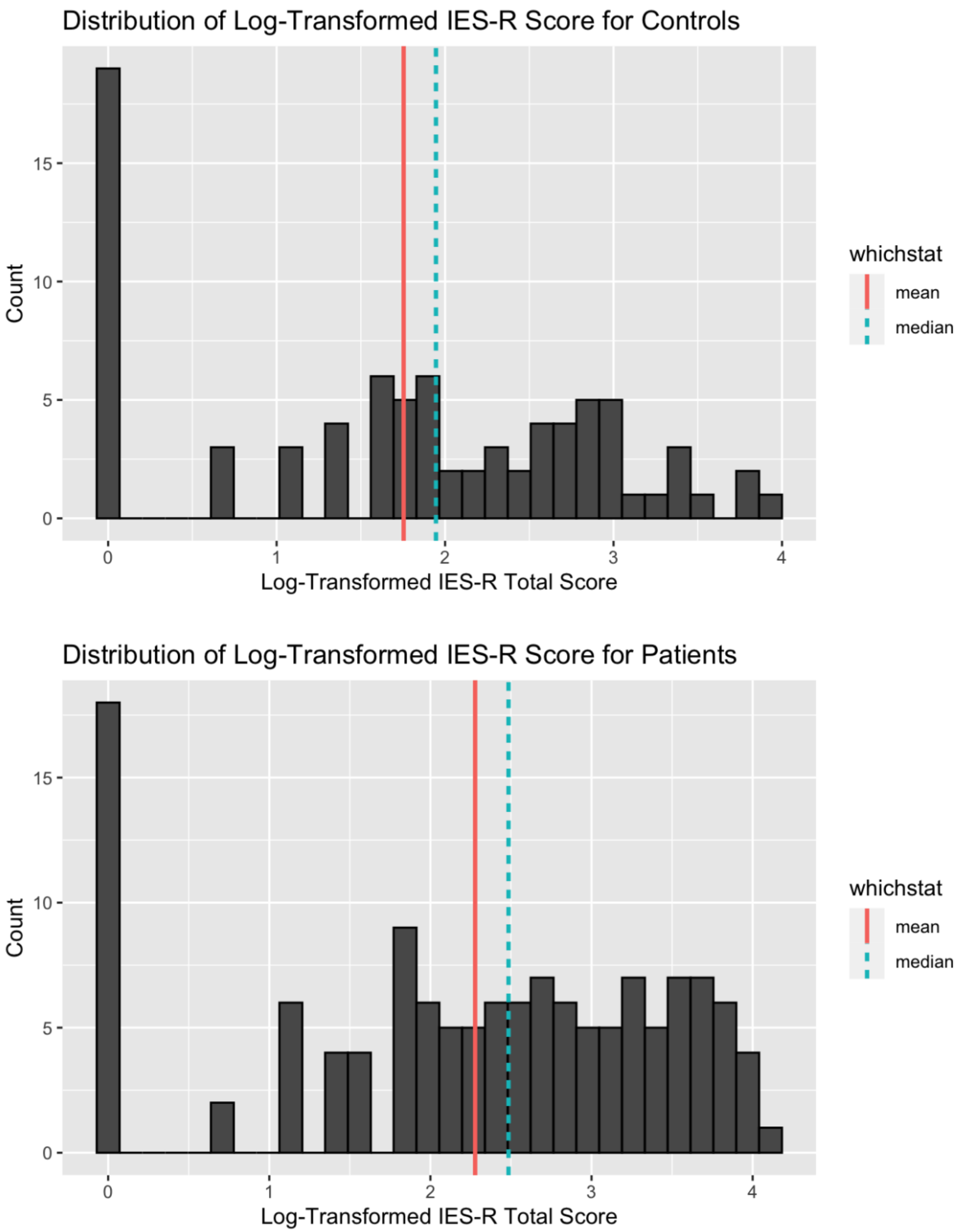


Supplementary Figure 2: Histogram of IES-R log-transformed score distribution. Among controls, mean IES-R is 1.75 (SD=1.19) and median is 1.95 (*IQR*=0.69-2.76). Among patients, mean score is 2.28 (SD=1.22) and median is 2.48 (*IQR*=1.61-3.28).
