## Supplemental Tables for "Pandemic-Related Post-traumatic Stress Symptomatology in COVID-19 Patients with and without Post-COVID Conditions"

*Table S1: Impact of Events Scale subscale scores*

| **Subscale** | **Controls (N=82)** | **Patients (N=131)** | **Mean Difference *[95% CI]*** | ***P* value** | **Cohen's *d* *[95% CI]*** | **PASC (N=68)** | **No PASC (N=63)** | **Mean Difference *[95% CI]*** | ***P* value** | **Cohen's *d* *[95% CI]*** |
|---|---|---|---|---|---|---|---|---|---|---|
| Median IES-R Intrusion Subscale *(IQR)* | 0.25 *(0.0-0.75)* | 0.50 *(0.13-1.25)* | 0.26 *[0.06, 0.46]* | 0.01 | 0.43 *[0.15, 0.71]* | 16.5 *(6.75-33)* | 6 *(2-15)* | 0.29 *[0.03, 0.56]* | 0.031 | 0.56 *[0.21, 0.91]* |
| Median IES-R Avoidance Subscale *(IQR)* | 0.25 *(0.0-0.63)* | 0.50 *(0.13-1.25)* | 0.22 *[0.05, 0.39]* | 0.013 | 0.42 *[0.14, 0.69]* | 0.81 *(0.25-1.66)* | 0.38 *(0.0-0.75)* | 0.30 *[0.07, 0.54]* | 0.013 | 0.62 *[0.27, 0.97]* |
| Median IES-R Hyperarousal Subscale *(IQR)* | 0.17 *(0.0-0.46)* | 0.33 *(0.0-1.0)* | 0.23 *[0.07, 0.39]* | 0.005 | 0.48 *[0.20, 0.76]* | 0.58 *(0.29-1.33)* | 0.16 *(0.0-0.5)* | 0.29 *[0.07, 0.51]* | 0.013 | 0.68 *[0.33, 1.03]* |

Table S2: Covariate coefficients for study glm models

| | **Patients/Controls** | | **PASC/No PASC** | |
|---|---|---|---|---|
| **Variable** | Estimate [*95% CI]* | p-value | Estimate [*95% CI]* | p-value |
| **Group** | 5.22 *[1.67,8.77]* | 0.004 | 6.49 *[1.70, 11.2])* | 0.009 |
| **Age** | -0.06 *[-0.20, 0.07]* | 0.362 | -0.17 *[-0.35, 0.04]* | 0.073 |
| **Gender:Male** | -1.25 *[-4.70, 2.19]* | 0.476 | 0.81 *[-3.91, 5.54]* | 0.736 |
| **Race:Black** | 1.97 *[-5.75, 9.70]* | 0.617 | 11.14 *[-0.53, 22.81]* | 0.064 |
| **Race:Other** | 0.75 *[-8.93, 10.42]* | 0.88 | 16.4 *[1.99, 30.81]* | 0.028 |
| **Race:White** | -3.60 *[-9.82, 2.62]* | 0.258 | 4.39 *[-4.91, 13.69]* | 0.357 |
| **MH tx hx** | 3.97 *[0.16, 7.79]* | 0.042 | 3.11 *[-2.07, 8.29]* | 0.241 |
| **GAD-2 cutoff** | 19.47 *[13.44, 25.51]* | <0.001 | 18.49 *[11.62, 25.37]* | <0.001 |

Table S3: Example IES-R open field free responses

| **Group** | **Free Response** |
|---|---|
| Patient | *Extreme exhaustion, Hallucination, and profusely sweating.* |
| Patient | *Learning a friend's brother suddenly died from Covid. He was in the hospital for four days with Covid, on Tuesday he was supposed to be released, but an hour before release his heart suddenly stopped. Doctors believe a blood clot resulting from him having Covid is the reason for his death.* |
| Patient | *The only time I was stressed was when I thought that me being positive could cause my grandma to get sick.* |
| Patient | *While I was sick, the most distressing thing was having my LFT's be at over 800. I was jaundiced and worried about going into liver failure. That was the most intense part of being sick and something I thought about in the hospital briefly but something I think about frequently now. It was, and still is, compounded by all of the uncertainty around COVID and what it does to the body in the long run…* |
| Patient | *Isolation* |
| Patient | *When I tested positive for COVID* |
| Patient | *Difficult question...most stressful has been the impact of not seeing family and friends.* |
| Patient | *Having on-going symptoms post a mild case of Covid* |
| Patient | *Being sick with COVID was one of the most serious illness I have had. In addition to being physically sick, the fear factor of getting an illness that has killed millions was very real and fore most on my mind. I kept on imagining all kinds of terrible scenarios, even my death. I even made my will and told everyone how I want things handled, if I am gone. However, as I am generally a positive person, part of me was determined to get well as well.* |
| Patient | *Not applicable - I don't have COVID related traumatic experiences* |
| Control | *Shortly after lockdown I had a panic attack. The only way to calm myself down was to lay on the floor. My family is in [country] and I miss them very much.* |

| | |
|---|---|
| Control | *Always wearing a mask, always getting your temperature taken.* |
| Control | *Very minimal trauma due to COVID-19, only inconveniences like partial shutdowns, etc.* |
| Control | *Caring for my mother when she was sick with covid. Dropping her off at the hospital and not knowing if that was the last time I'd see her. Talking with her doctor every day and not seeing any improvement, hearing how bad all the tests came back. Then, after she received two negative covid tests, the hospital discharging her because they were going covid-only and releasing her with a high fever and no diagnosis. Being solely responsible for her well being in the week that followed without any medical training. Not sleeping so I could check on her throughout the night to make sure she was still breathing…* |
| Control | *My company & job being shut down indefinitely* |
| Control | *Don't know* |
| Control | *My most stressful experience related to COVID-19 is hearing about so many people who have died alone in the hospital.* |
| Control | *The most stressful experience for me re COVID-19 was getting symptoms that seemed to suggest I had COVID last March, before tests were available. There was so little known, and no place to go. My head throbbed, my throat was sore, and my breathing was labored off and on for about a month. My PCP had me quarantine at home. I never had fever or cough, but I did suddenly develop severe right flank pain that sent me to urgent care, then to the ER, for fear it was my kidneys. Turned out I did not have Covid at all, but living with what my doctors thought was covid for that one month, reading daily stories in the news of people whose mild symptoms suddenly got worse and they died overnight, was frightening…* |
| Control | *5 of my family members got sick from COVID-19. I was really worried for my grandmom as I have seen many elderly die from this disease.* |
| Control | *Working on a covid unit, having to care for dying hospice covid patients, when originally no visitors were allowed in the hospital. Holding hands with a dying patient, watching them take their last breath, with no family physically present but on a zoom call, all crying.* |
